# Comparison of the Cholesterol, High-Density Lipoprotein, and Glucose (CHG) Index, Atherogenic Index of Plasma (AIP), and Triglyceride-Glucose (TyG) Index in Predicting the Risk of New-Onset Hypertension Among Prehypertensive Individuals: A Cohort Study

**DOI:** 10.1101/2025.05.05.25327038

**Authors:** Mengmeng Wang, Zhankui Du, Tianqi Teng, Jiachao Xu, Zihan Dong, Qingying Jiao, Ning Zhang, Haichu Yu

## Abstract

**Background:** Early identification of individuals at high risk for hypertension development is crucial for implementing timely preventive strategies. Metabolic indices such as the cholesterol-glucose (CHG) index, atherogenic index of plasma (AIP), and triglyceride-glucose (TyG) index have emerged as potential biomarkers for metabolic and cardiovascular disorders. However, their comparative predictive value for new-onset hypertension in prehypertensive individuals remains unclear.

**Methods:** This prospective cohort study utilized data from the China Health and Retirement Longitudinal Study (CHARLS), including 2,859 adults with prehypertension followed from 2011 to 2015. Participants were stratified based on progression to incident hypertension. Baseline characteristics and metabolic indices were evaluated. Multivariable logistic regression models, restricted cubic spline (RCS) analyses, receiver operating characteristic (ROC) curves, and subgroup analyses were conducted to assess the associations between CHG, AIP, and TyG indices and the risk of developing hypertension.

**Results:** During the 4-year follow-up, 31.34% (896/2,859) of participants developed new-onset hypertension. All three metabolic indices were independently associated with an increased risk of hypertension after multivariable adjustment. The CHG index demonstrated the strongest association (odds ratio [OR]: 1.96, 95% confidence interval [CI]: 1.45–2.66, P < 0.001), followed by the TyG index (OR: 1.31, 95% CI: 1.07–1.60, P = 0.010). RCS analysis revealed a significant nonlinear relationship between the CHG index and hypertension risk (P for nonlinear = 0.042), whereas AIP and TyG showed linear trends. ROC analysis indicated that the CHG index had the highest discriminatory ability for predicting hypertension (fully adjusted area under the curve [AUC] = 0.7010), outperforming both AIP (AUC = 0.6997) and TyG (AUC = 0.6980). Subgroup analyses showed that the association between the CHG index and incident hypertension was significantly stronger among individuals with lower educational attainment (illiterate), those aged 60–70 or ≥70 years, and widowed individuals (P for interaction < 0.05).

**Conclusion:** Among prehypertensive individuals, higher baseline levels of CHG, AIP, and TyG indices are significantly associated with an increased risk of developing hypertension. The CHG index demonstrates superior predictive performance and may serve as a valuable tool for early risk stratification and targeted intervention in clinical practice.

## 1. Introduction

Prehypertension, defined as systolic blood pressure (SBP) ranging from 120 to 139 mmHg or diastolic blood pressure (DBP) between 80 and 89 mmHg, represents a critical transitional stage in cardiovascular disease (CVD) progression^1^. This condition has gained increasing recognition as a key intervention target in preventive cardiology due to its substantial global burden and clinical implications^2^. Epidemiological evidence reveals three particularly concerning characteristics of prehypertension: (1) significant demographic variations in prevalence, with rates of 30-35% in East Asian populations (China 34.5%, South Korea 31.6%) compared to 40-50% in Western population^3–5^; (2) alarming progression rates, including a lifetime risk approaching 90% in individuals aged ≥55 years and 3-year conversion rates reaching 44%^6^; and (3) direct associations with a 1.5- to 2-fold increased risk of cardiovascular events independent of progression to overt hypertension^7^. Evidence from systematic reviews of prospective cohort studies demonstrates that each 10-mmHg reduction in SBP is associated with a significant 33% reduction in stroke risk among individuals aged 60-79 years^8^. Blood pressure is strongly linked to vascular mortality, with significant associations observed even at levels as low as 115/75 mmHg^9^. According to the World Health Organization, effective blood pressure control interventions could potentially save approximately US$1.2 trillion in global healthcare expenditures by 2030^2^. Therefore, improving blood pressure management in the prehypertensive population has become an urgent public health priority.

Building upon this clinical imperative, recent advances in metabolic biomarkers have provided new opportunities for hypertension risk prediction. Among these markers, the atherogenic index of plasma (AIP = log[triglycerides (TG, mg/dL)/high-density lipoprotein cholesterol (HDL-C, mg/dL)]) has emerged as a particularly promising indicator^10^. First proposed by Dobiásová in 2000, AIP quantifies the ratio of TG to HDL-C, providing a sensitive measure of dyslipidemia and atherogenic propensity^10^. The clinical relevance of AIP extends beyond its well-established associations with coronary artery disease and ischemic stroke^11,12^; mounting evidence suggests it may surpass traditional lipid parameters in predicting cardiovascular risk^13^. Previous studies have reported a positive correlation between AIP and prehypertension or hypertension^14,15^. Another study showed that AIP only demonstrated predictive value for hypertension after adjusting for age and C-reactive protein^16^. However, the predictive utility of AIP for hypertension remains controversial, with studies reporting conflicting results regarding its population-specific performance^14–17^. This heterogeneity highlights important gaps in our understanding of AIP’s clinical applicability.

Parallel developments in insulin resistance assessment have identified the triglyceride-glucose index (TyG, calculated as ln[fasting TG (mg/dL) × fasting blood glucose (FBG, mg/dL)/2]) as another valuable predictive tool^18^. Since its introduction by Simental-Mendía et al. in 2008, TyG index has gained recognition as a robust surrogate marker of insulin resistance that captures essential features of glucose and lipid metabolism dysregulation^18^. A study involving 32,124 adults with normal glucose levels showed that when comparing the highest versus the lowest TyG index categories, there was a significant association with both prehypertension and hypertension, with corresponding odds ratios of 1.795 (95% confidence interval [CI]: 1.638–1.968) and 2.439 (95% CI: 2.205–2.698), respectively^19^. However, inconsistent findings across community-based studies and insufficient data regarding its relationship with ambulatory blood pressure parameters suggest that TyG index alone may not provide comprehensive risk assessment^20–23^.

Most recently, the cholesterol-high-density lipoprotein-glucose index (CHG index = ln[(total cholesterol (TC, mg/dL) × FBG (mg/dL))/(2 × HDL-C (mg/dL))]) has emerged as a novel integrated metabolic marker with potential applications in cardiovascular risk prediction^24^. Developed by Mansoori’s team in 2024, this innovative index represents a significant conceptual advance by simultaneously evaluating three critical metabolic pathways: cholesterol metabolism, reverse cholesterol transport, and glucose homeostasis^24^. While preliminary studies have demonstrated CHG’s superior diagnostic performance compared to the TyG index in diabetes screening (area under the curve [AUC]: 0.864 vs 0.825), reflecting its more comprehensive incorporation of metabolic syndrome pathophysiology, its clinical utility extends beyond this single application^24^. Notably, despite its strong theoretical foundation and practical advantages through the use of routinely measured parameters, crucial gaps remain in our understanding of CHG index’s predictive value for hypertension development, particularly in prehypertensive populations.

To address these critical knowledge gaps, we conducted the first comprehensive cohort study to systematically evaluate and compare all three metabolic indices (CHG, AIP, and TyG) for hypertension prediction in prehypertensive adults using data from the China Health and Retirement Longitudinal Study (CHARLS). Our four-year prospective investigation has three primary objectives: (1) to establish the predictive validity of the novel CHG index for incident hypertension in this at-risk population; (2) to perform head-to-head comparisons of its performance against established indices (AIP and TyG); and (3) to assess the clinical utility of these indices for precision risk stratification in primary prevention settings.

## 2. Methods

### 2.1 Research Design

This study employed a retrospective cohort design to systematically evaluate the associations between baseline metabolic indices—CHG, AIP, and TyG index—and the risk of incident hypertension. Using longitudinal data from the CHARLS, we aimed to assess the predictive value of these metabolic markers for hypertension development and compare their relative performance. Specifically, both unadjusted and multivariable-adjusted logistic regression models were applied to examine the independent relationships between each index and new-onset hypertension, while controlling for potential confounding factors. Restricted cubic spline (RCS) analysis was further conducted to explore potential nonlinear associations. Additionally, the predictive performance of each index was compared using receiver operating characteristic (ROC) curve analysis across different follow-up periods.

### 2.2 Data Source and Study Population

Data for this study were obtained from the CHARLS, a nationally representative, ongoing cohort study launched in 2011 to investigate health and aging among Chinese adults aged 45 years and older (http://charls.pku.edu.cn/)^25^. The study employed a multistage stratified random sampling design covering 150 counties across 28 provinces, with an initial response rate of 80.5%^26^. This approach minimized selection bias and ensured broad demographic representation. Baseline data were collected in 2011, with follow-up assessments conducted in 2015. From the original sample, participants were selected based on the following inclusion criteria: (1) availability of baseline biochemical measurements; (2) complete household roster information; (3) presence of prehypertension at baseline, defined as SBP between 120 and 139 mmHg or DBP between 80 and 89 mmHg; (4) valid blood pressure measurements at the 2015 follow-up; and (5) complete data on CHG, AIP, and TyG indices. After applying these criteria, a total of 2,859 individuals were included in the final analysis (Figure 1).

**Fig. 1.**
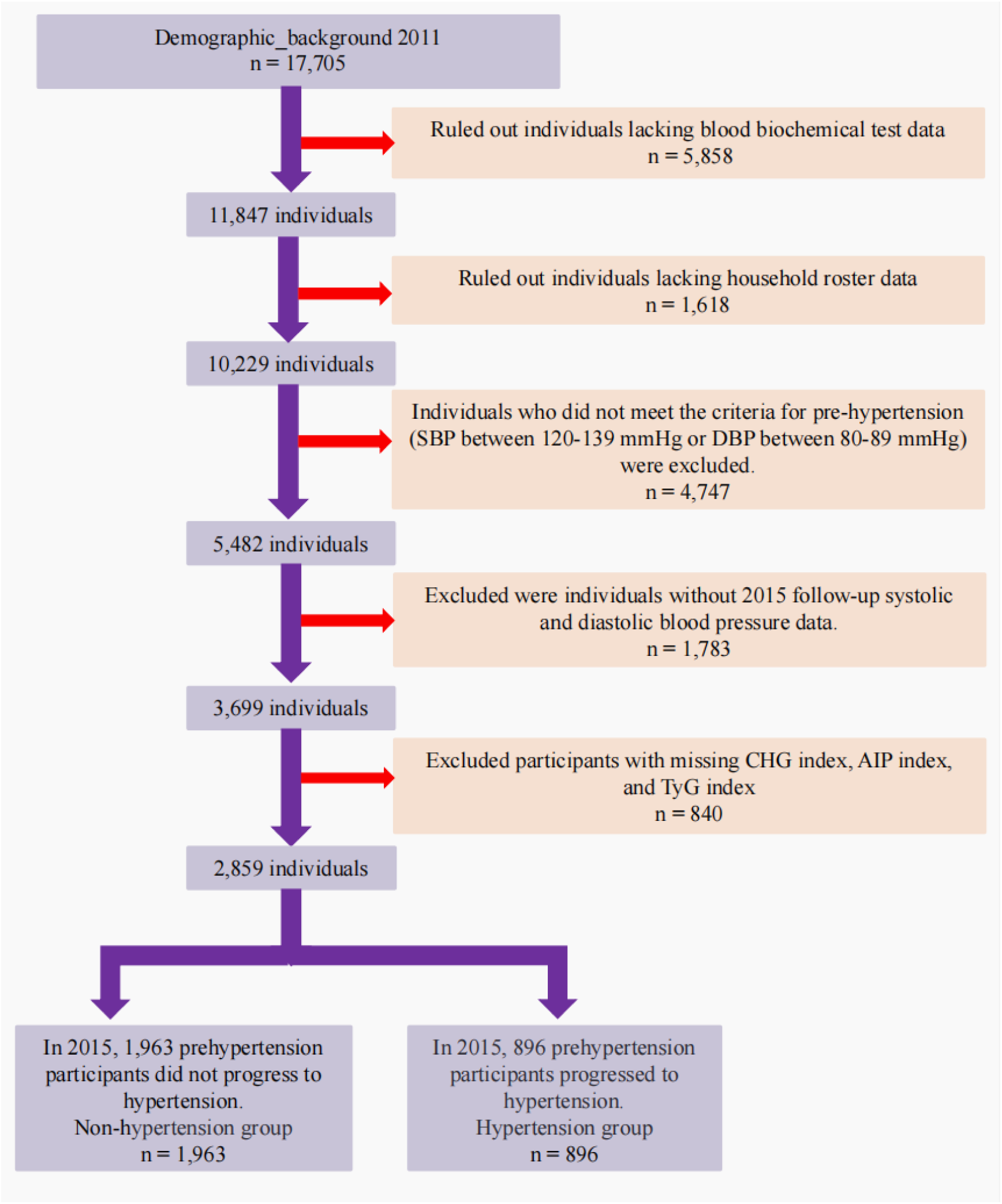
The flowchart of this study

The CHARLS study was approved by the Institutional Review Board at Peking University (IRB00001052-11015), in accordance with the principles outlined in the Declaration of Helsinki. All participants provided written informed consent prior to enrollment. This study followed the STROBE guidelines for reporting observational research, ensuring methodological rigor and transparency throughout the study design, implementation, and reporting.

### 2.3 Evaluation of CHG index, AIP index and TyG index

For a comprehensive evaluation of the CHG index, AIP index, and TyG index, this study utilized data from the clinical laboratory of Beijing You’an Hospital Affiliated with Capital Medical University. Measurements of TG and FBG levels were performed using enzymatic colorimetric techniques on frozen plasma or whole blood samples, with a coefficient of variation maintained below 2% to ensure data accuracy and reliability^27^. The CHG index, integrating TC, HDL-C), and glucose levels, was calculated using the formula: CHG index = ln[(TC (mg/dL) × FBG (mg/dL))/(2 × HDL-C (mg/dL))], providing a comprehensive assessment of an individual’s metabolic health status^24^. The AIP was defined as the logarithm of the ratio of plasma TG to HDL-C: AIP = log(TG (mg/dL)/HDL-C (mg/dL)), which not only reflects changes in lipid profiles but also indirectly indicates the degree of insulin resistance^10^. The TyG index, serving as a valid surrogate marker for insulin resistance, was calculated using the formula: TyG index = ln[(TG (mg/dL) × FBG (mg/dL))/2], offering a simple yet accurate method to assess individual insulin sensitivity by combining fasting glucose and triglyceride levels^18^. For analytical purposes, each index value was categorized into tertiles: T1 (lowest third), T2 (middle third), and T3 (highest third), with T1 serving as the reference group. This stratification approach facilitates an in-depth investigation of the relationship between different metabolic levels and incident hypertension, while revealing the potential role of these indices in predicting disease risk.

### 2.4 Assessment of Incident Hypertension

This study aimed to identify the development of hypertension during a four-year follow-up (2011–2015) among participants who were classified as prehypertensive at baseline, defined as having SBP of 120–139 mmHg or DBP of 80–89 mmHg in 2011. All participants were followed until the end of the study period, ensuring completeness of outcome data and reliability of event ascertainment.

### 2.5 Data Collection in the Cohort Study

To comprehensively investigate risk factors for incident hypertension among individuals with prehypertension, a broad range of data was collected. These included demographic characteristics such as age, gender, education level, marital status, and residential area; lifestyle factors including smoking and alcohol consumption; physical measurements such as body mass index (BMI), waist circumference, baseline SBP, and DBP; and laboratory parameters including uric acid (mg/dL), creatinine (mg/dL), cystatin C (mg/dL), and blood urea nitrogen (BUN, mg/dL), white blood cell count (WBC × 10⁹/L), platelet count (PLT × 10⁹/L), hemoglobin (HGB, g/dL), TG (mg/dL), FBG (mg/dL), TC (mg/dL), glycated hemoglobin (HbA1c), and low-density lipoprotein cholesterol (LDL-C, mg/dL). Comorbid conditions such as CVD and diabetes mellitus (DM), as well as mental health indicators including depressive symptoms and life satisfaction, were also recorded. Participant characteristics were classified according to multiple variables. Marital status was categorized as divorced, married, unmarried, or widowed. Education level was grouped into four categories: illiterate, primary school, middle school, and high school or above. Age was stratified into <50 years, 50–60 years, 60–70 years, and ≥70 years. Residential area was classified as urban or rural. Smoking status was defined as non-smoker, former smoker, or current smoker. Alcohol consumption frequency was categorized as non-drinker, less than once per month, or more than once per month. BMI was divided into four groups: <18.5, 18.5–24, 24–28, and ≥28. The presence or absence of CVD and DM was documented, and depressive symptoms were classified as absent or present. Life satisfaction was further categorized into five levels: 0, 1, 2, 3, and 4, representing increasing degrees of satisfaction.

### 2.6 Definition and Assessment of Comorbidities

In the CHARLS database, comorbid conditions were determined based on participants’ self-reported medical histories and biochemical test results. CVD was defined as a participant reporting a history of heart disease or stroke. If a participant reported either condition, they were considered to have CVD; otherwise, they were classified as not having CVD. DM was diagnosed using HbA1c levels, self-reported DM history, and the use of antidiabetic medications. Participants with an HbA1c level below 6.5%, no reported history of DM, and no use of antidiabetic medications were classified as not having DM. Conversely, participants meeting any of these criteria (HbA1c ≥ 6.5%, self-reported DM, or use of antidiabetic medications) were classified as having DM. Mental health status was assessed using the Center for Epidemiologic Studies Depression Scale (CES-D), which measures the severity of depressive symptoms. A CES-D score of 10 or higher was considered indicative of depression^28^.

### 2.8 Statistical Analysis

Baseline characteristics of participants were summarized using descriptive statistics and stratified by the occurrence of incident hypertension during follow-up. The normality of continuous variables was assessed using the Shapiro-Wilk test. Continuous variables were presented as mean ± standard deviation (SD) for normally distributed data or median (interquartile range, IQR) for non-normally distributed data. Categorical variables were expressed as frequency (percentage). Comparisons between groups were performed using Fisher’s exact test or chi-square test for categorical variables, and Mann-Whitney U test for continuous variables. When comparing more than three groups, Kruskal-Wallis rank-sum test was applied for continuous variables and Pearson chi-square test for categorical variables.

To evaluate the associations between baseline CHG index, AIP index, TyG index, and the risk of incident hypertension, univariable and multivariable logistic regression models were employed. Covariates included in the multivariable models were selected based on clinical relevance and statistical significance. Each metabolic index was analyzed both as a continuous variable and as a categorical variable stratified into tertiles (T1–T3), including CHG (T1: 3.6857–5.1550, T2: 5.1550–5.5163, T3: 5.5163–7.9643), AIP (T1: −1.1385–0.4696, T2: 0.4696–1.1609, T3: 1.1609–4.3291), and TyG (T1: 6.2845–8.3928, T2: 8.3928–8.9502, T3: 8.9502–12.2656). Four logistic regression models were constructed to evaluate their associations with incident hypertension, including an unadjusted model (Model 1), a model adjusted for demographic and lifestyle factors along with baseline blood pressure (Model 2), a model further adjusted for laboratory biomarkers (Model 3), and a fully adjusted model also accounting for comorbidities such as CVD, DM, and depressive symptoms (Model 4).

The discriminatory ability of each metabolic index in predicting incident hypertension was evaluated using ROC curve analysis, with comparisons based on AUC. To explore potential nonlinear relationships between the indices and the risk of new-onset hypertension, RCS models were fitted based on logistic regression, with the optimal number of knots selected using the Bayesian information criterion (BIC). The same set of covariates used in the logistic regression models was applied in the RCS analyses.

Subgroup analyses were conducted to assess the consistency of observed associations across different population strata. Stratification variables included age, sex, marital status, education level, residence area, alcohol consumption, smoking status, BMI, presence of DM, CVD, depression, and life satisfaction. Potential effect modification was evaluated by introducing interaction terms into the models. All statistical analyses were performed using R software (version 4.4.1). Two-sided p-values < 0.05 were considered statistically significant.

## 3. Results

### 3.1 Characteristics of New-Onset Hypertension in Prehypertensive Participants After 4 Years

Table 1 presents the demographic and clinical characteristics of prehypertensive participants stratified by whether they progressed to incident hypertension over the period from 2011 to 2015. The mean age of the study cohort was 59.80 years, with 44.46% being female. Among the total of 2,859 participants, 896 (31.34%) were diagnosed with new-onset hypertension by the end of the follow-up. Compared to those who did not develop hypertension, participants who eventually developed hypertension exhibited several significant differences: they were older on average, with a higher proportion in the 60–70 age group; a greater percentage were male; and their BMI values were generally higher, particularly in the ranges of 24–28 and ≥28. Notably, the new-onset hypertension group was more likely to reside in rural areas. Further observations revealed that waist circumference, SBP, DBP, uric acid levels, HGB, TG, FBG, BUN, creatinine, as well as CHG index, AIP index, and TyG index were all higher in the new-onset hypertension group compared to the non-progressing group. Additionally, the prevalence of CVD and DM was also higher in the new-onset hypertension group. However, there were no significant differences between the two groups in terms of alcohol consumption, education level, PLT, LDL-C, TC, HbA1c, cystatin C, depression status, and life satisfaction (all P-values > 0.05).

**Table 1.** Demographic and clinical attributes of prehypertensive participants stratified by the development of new-onset hypertension.

| Characteristic | Overall<br>N = 2,859<sup>1</sup> | Non-hypertension<br>N = 1,963<sup>1</sup> | Hypertension<br>N = 896<sup>1</sup> | P-value<sup>2</sup> |
| --- | --- | --- | --- | --- |
| Age, years | 59.80 (9.58) | 59.20 (9.38) | 61.12 (9.88) | <0.001 |
| Age group, N(%) |  |  |  | <0.001 |
| <50 | 574 (20.08%) | 420 (21.40%) | 154 (17.19%) |  |
| 50-60 | 1,011 (35.36%) | 738 (37.60%) | 273 (30.47%) |  |
| 60-70 | 888 (31.06%) | 565 (28.78%) | 323 (36.05%) |  |
| ≥70 | 386 (13.50%) | 240 (12.23%) | 146 (16.29%) |  |
| Gender, N(%) |  |  |  | 0.005 |
| Female | 1,275 (44.66%) | 910 (46.45%) | 365 (40.74%) |  |
| Male | 1,580 (55.34%) | 1,049 (53.55%) | 531 (59.26%) |  |
| Education, N(%) |  |  |  | >0.900 |
| High school+ | 319 (11.16%) | 216 (11.00%) | 103 (11.50%) |  |
| Illiterate | 615 (21.51%) | 429 (21.85%) | 186 (20.76%) |  |
| Middle school | 637 (22.28%) | 435 (22.16%) | 202 (22.54%) |  |
| Primary | 1,288 (45.05%) | 883 (44.98%) | 405 (45.20%) |  |
| Marital, N(%) |  |  |  | 0.002 |
| Divorced | 54 (1.89%) | 42 (2.14%) | 12 (1.34%) |  |
| Married | 2,310 (80.80%) | 1,615 (82.27%) | 695 (77.57%) |  |
| Unmarried | 24 (0.84%) | 15 (0.76%) | 9 (1.00%) |  |
| Widowed | 471 (16.47%) | 291 (14.82%) | 180 (20.09%) |  |
| Residential area, N(%) |  |  |  | 0.015 |
| Urban | 230 (8.04%) | 141 (7.18%) | 89 (9.93%) |  |
| Rural | 2,629 (91.96%) | 1,822 (92.82%) | 807 (90.07%) |  |
| Smoking, N(%) |  |  |  | <0.001 |
| Ex-smoker | 309 (10.83%) | 183 (9.35%) | 126 (14.06%) |  |
| Non-smoker | 1,603 (56.19%) | 1,112 (56.82%) | 491 (54.80%) |  |
| Smoker | 941 (32.98%) | 662 (33.83%) | 279 (31.14%) |  |
| Drinking, N(%) |  |  |  | 0.400 |
| Drink but less than once a month | 216 (7.57%) | 149 (7.61%) | 67 (7.48%) |  |
| Drink more than once a month | 796 (27.88%) | 531 (27.11%) | 265 (29.58%) |  |
| None of these | 1,843 (64.55%) | 1,279 (65.29%) | 564 (62.95%) |  |
| BMI, kg/m<sup>2</sup> | 23.73 (3.99) | 23.46 (4.04) | 24.31 (3.82) | <0.001 |
| BMI group, N(%) |  |  |  | <0.001 |
| <18.5 kg/m<sup>2</sup> | 180 (6.30%) | 140 (7.13%) | 40 (4.46%) |  |
| ≥28 kg/m<sup>2</sup> | 356 (12.45%) | 216 (11.00%) | 140 (15.63%) |  |
| 18.5-24 kg/m<sup>2</sup> | 1,466 (51.28%) | 1,062 (54.10%) | 404 (45.09%) |  |
| 24-28 kg/m<sup>2</sup> | 857 (29.98%) | 545 (27.76%) | 312 (34.82%) |  |
| Waist circumference,cm | 84.97 (12.34) | 84.02 (12.43) | 87.06 (11.88) | <0.001 |
| SBP_baseline,mmHg | 128.96 (6.12) | 128.17 (6.01) | 130.68 (6.01) | <0.001 |
| DBP_baseline,mmHg | 75.94 (7.15) | 75.59 (7.03) | 76.72 (7.36) | <0.001 |
| Uric acid, mg/dL | 270.15 (77.03) | 266.46 (73.45) | 278.24 (83.80) | <0.001 |
| WBC, x10<sup>9</sup>/L | 6.37 (1.88) | 6.42 (1.91) | 6.26 (1.82) | 0.013 |
| HGB, g/dL | 14.63 (2.21) | 14.48 (2.06) | 14.95 (2.49) | <0.001 |
| PLT, x10<sup>9</sup>/L | 213.62 (74.74) | 214.78 (78.20) | 211.09 (66.50) | >0.900 |
| TC, mg/dL | 194.06 (37.44) | 193.68 (37.23) | 194.88 (37.91) | 0.200 |
| TG, mg/dL | 136.17 (101.06) | 127.99 (88.62) | 154.10 (122.17) | <0.001 |
| HDL, mg/dL | 50.27 (15.26) | 51.48 (15.20) | 47.62 (15.07) | <0.001 |
| LDL, mg/dL | 116.73 (35.49) | 117.38 (34.95) | 115.31 (36.63) | 0.600 |
| FBG, mg/dL | 112.31 (36.15) | 109.97 (32.48) | 117.42 (42.68) | <0.001 |
| HbA1c, % | 5.32 (0.83) | 5.30 (0.79) | 5.35 (0.93) | 0.700 |
| CVD, N(%) |  |  |  | <0.001 |
| No | 2,466 (86.25%) | 1,724 (87.82%) | 742 (82.81%) |  |
| Yes | 393 (13.75%) | 239 (12.18%) | 154 (17.19%) |  |
| DM, N(%) |  |  |  | <0.001 |
| No | 2,593 (90.70%) | 1,811 (92.26%) | 782 (87.28%) |  |
| Yes | 266 (9.30%) | 152 (7.74%) | 114 (12.72%) |  |
| BUN, mg/dL | 15.67 (4.20) | 15.40 (3.99) | 16.27 (4.58) | <0.001 |
| Creatinine, mg/dL | 0.79 (0.18) | 0.78 (0.17) | 0.82 (0.20) | <0.001 |
| Cystatin_C, mg/dL | 1.00 (0.25) | 1.00 (0.24) | 1.01 (0.27) | 0.900 |
| Depression, N(%) |  |  |  | 0.500 |
| No | 1,824 (63.80%) | 1,261 (64.24%) | 563 (62.83%) |  |
| Yes | 1,035 (36.20%) | 702 (35.76%) | 333 (37.17%) |  |
| Life Satisfaction, N(%) |  |  |  | 0.100 |
| 0 | 68 (2.38%) | 56 (2.85%) | 12 (1.34%) |  |
| 1 | 387 (13.54%) | 273 (13.91%) | 114 (12.72%) |  |
| 2 | 1,812 (63.38%) | 1,239 (63.12%) | 573 (63.95%) |  |
| 3 | 561 (19.62%) | 374 (19.05%) | 187 (20.87%) |  |
| 4 | 31 (1.08%) | 21 (1.07%) | 10 (1.12%) |  |
| CHG index | 5.37 (0.47) | 5.33 (0.44) | 5.47 (0.50) | <0.001 |
| AIP index | 0.87 (0.78) | 0.80 (0.73) | 1.02 (0.84) | <0.001 |
| TyG index | 8.74 (0.67) | 8.68 (0.64) | 8.86 (0.72) | <0.001 |

<sup>1</sup>Mean (SD); n (%)
<sup>2</sup>Kruskal-Wallis rank sum test; Pearson's Chi-squared test

### 3.2 Baseline Characteristics of Participants Classified by Tertiles of CHG Index, AIP Index, and TyG Index

Table 2 and Supplementary Tables S1 and S2 present the baseline characteristics of participants classified by tertiles of the CHG index, AIP index, and TyG index. For participants in the highest tertile (T3) of the CHG index, several notable characteristics were observed: most participants were aged between 50 and 60 years; marital status was predominantly married; and educational attainment was mainly at the middle school level or above. Notably, although the majority resided in rural areas, the proportion of urban residents was higher in the T3 group compared to the lower tertiles. This group also exhibited significant differences in various health indicators: higher BMI values, particularly in the ranges of 24–28 and ≥28; larger waist circumference; elevated DBP; and increased levels of uric acid, WBC, HGB, PLT, TG, LDL-C, FBG, HbA1c,TC, and creatinine. Additionally, the prevalence of CVD and DM was higher in this group. However, no significant differences were observed among the tertiles in terms of gender distribution, BUN levels, depression status, and life satisfaction (all P-values > 0.05). For the AIP index, some unique characteristics were noted that differed from those observed for the CHG index: SBP and education level did not significantly differ across tertiles, and the sample was predominantly female. The T3 group of the AIP index showed higher levels of HbA1c. Similarly, analysis of the TyG index revealed differences within its tertiles: marital status, SBP, and PLT did not significantly differ across groups. Despite these differences, the analyses of both the AIP and TyG indices largely mirrored the trends observed with the CHG index, including higher levels of metabolic markers and a greater prevalence of CVD and DM in the highest tertile.

**Table 2.**
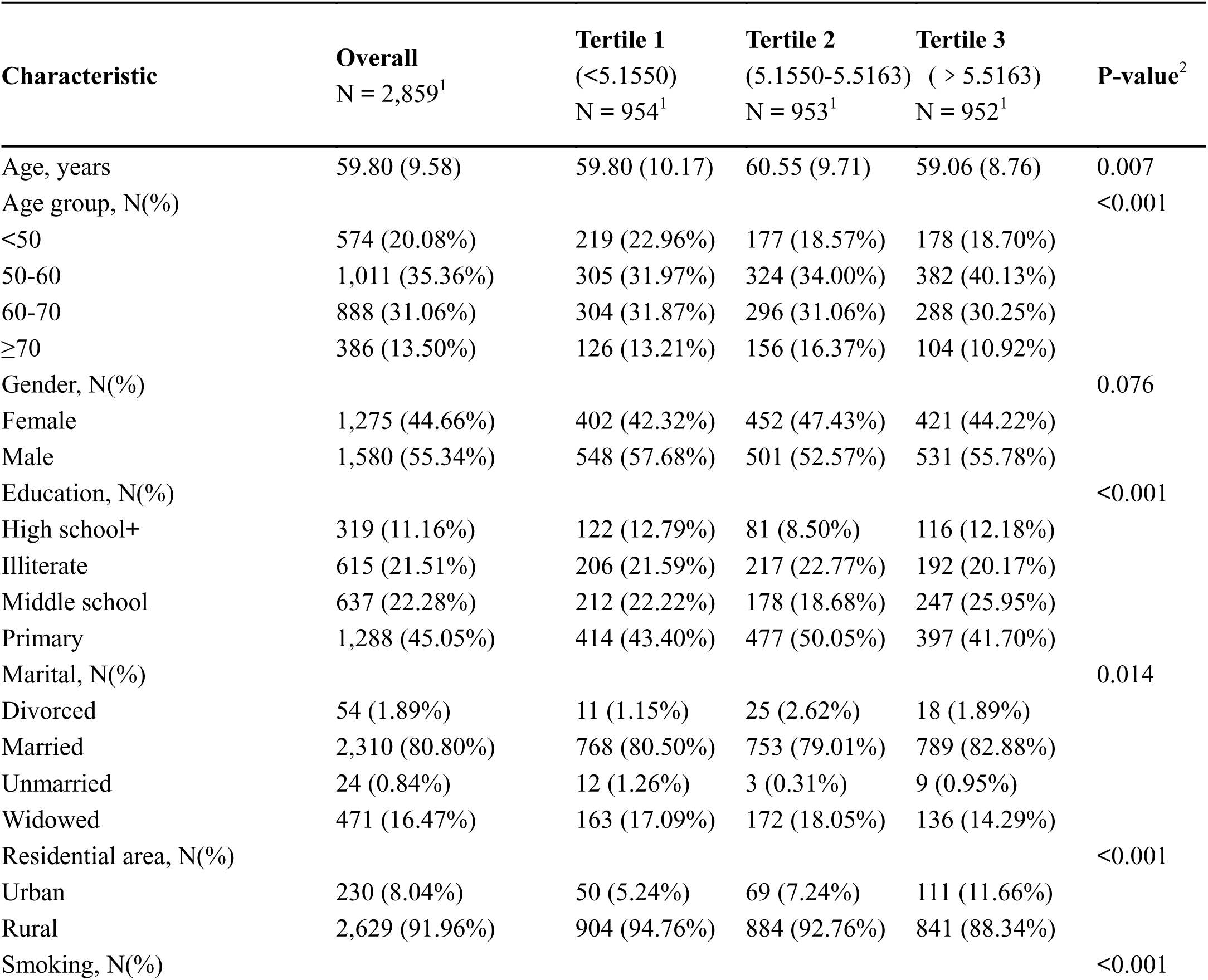
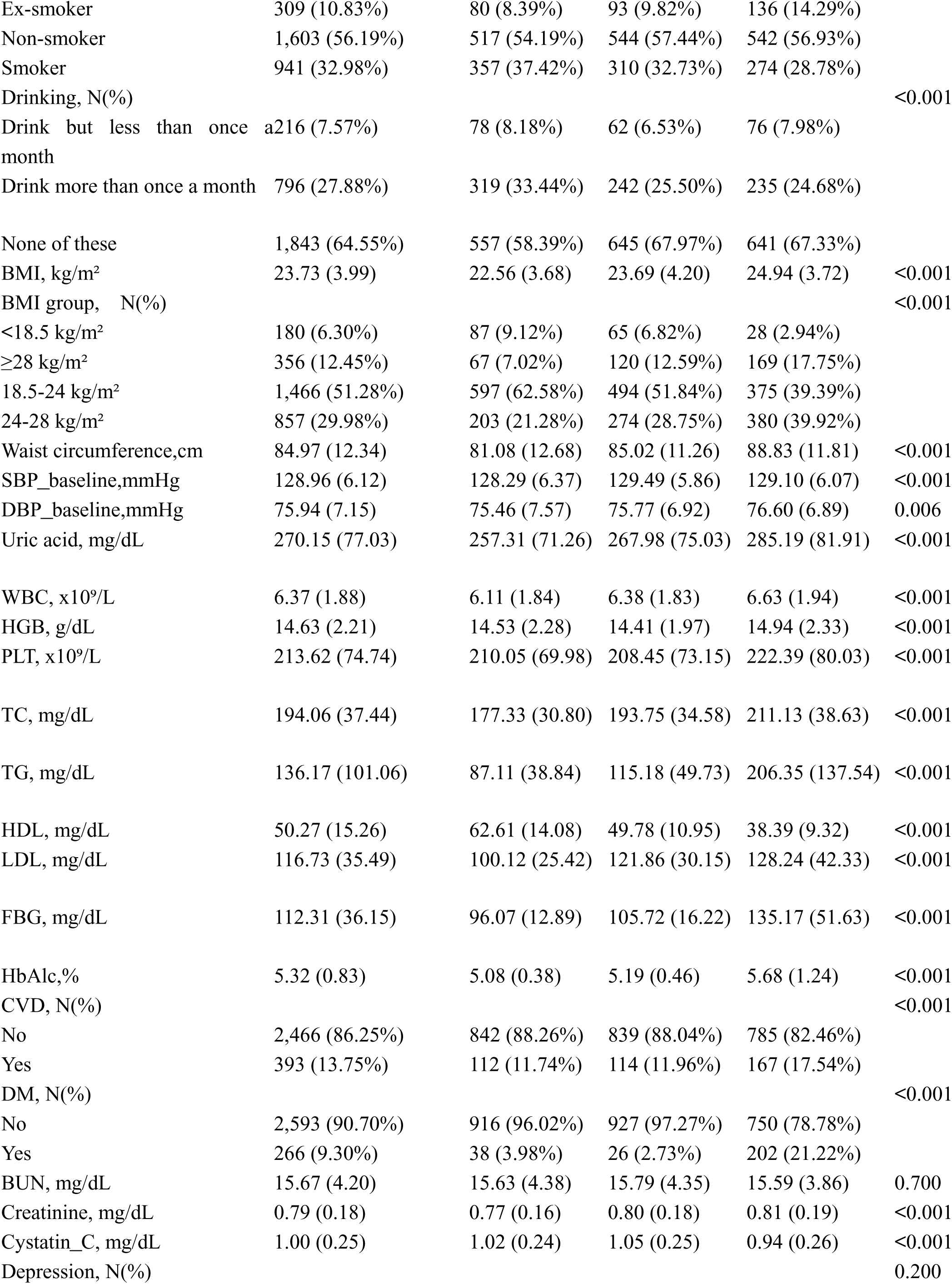
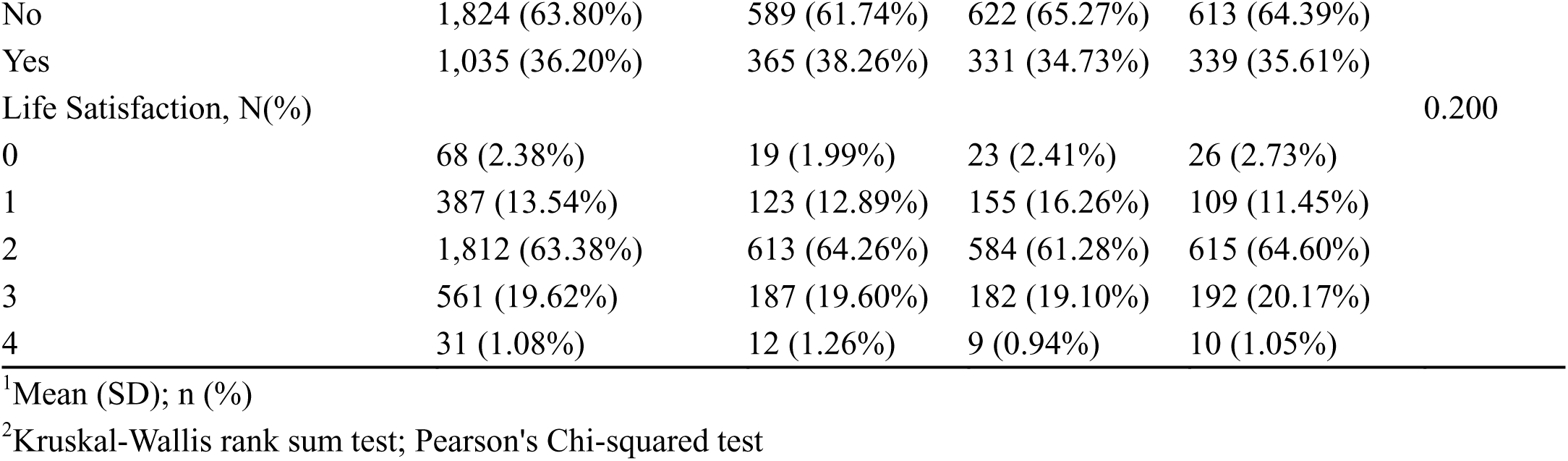
Baseline Attributes of Participants Divided into CHG Index Tertiles.

### 3.3 Logistic Regression Model Evaluation of the Association Between CHG Index, AIP Index, and TyG Index and New-Onset Hypertension

Table 3 presents the associations between the CHG index, AIP index, TyG index, and the risk of incident hypertension. Multivariable logistic regression analyses revealed significant positive relationships between each of these continuous metabolic indices and the occurrence of new-onset hypertension. In the fully adjusted Model 4, the CHG index demonstrated the strongest association, with a 96% increase in the risk of developing hypertension per unit increase (odds ratio [OR]: 1.96, 95% CI: 1.45–2.66, P < 0.001), followed by the TyG index (OR: 1.31, 95% CI: 1.07–1.60, P = 0.010). When these indices were categorized into tertiles, higher tertile groups (T2 and T3) were consistently associated with increased risks of incident hypertension across all four models for the CHG, AIP, and TyG indices. Specifically, in the fully adjusted Model 4, compared to the lowest tertile (T1), participants in the highest tertile (T3) of the CHG index had the greatest risk of developing hypertension (OR: 1.90, 95% CI: 1.45–2.50, P < 0.001), followed by those in the T3 group of the AIP index (OR: 1.51, 95% CI: 1.20–1.91, P < 0.001), and the TyG index (OR: 1.36, 95% CI: 1.05–1.76, P = 0.020). These findings indicate that elevated levels of the CHG, AIP, and TyG indices are independently associated with an increased risk of new-onset hypertension. These metabolic indices may therefore serve as valuable tools for risk stratification and early prediction of hypertension development.

**Table 3.**
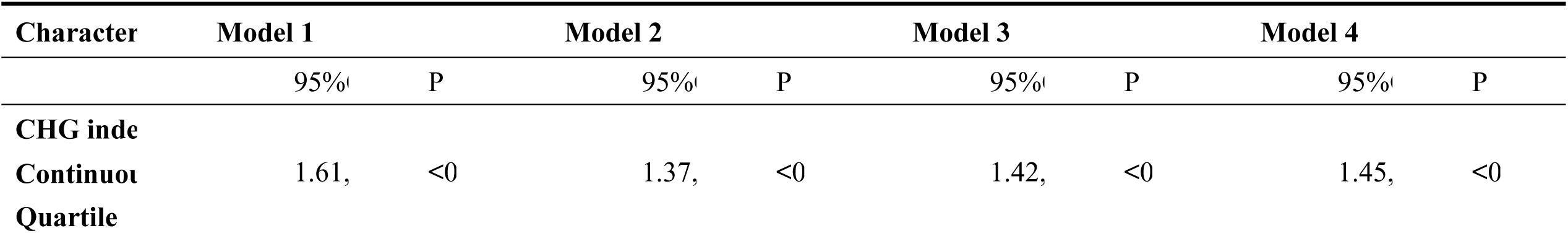
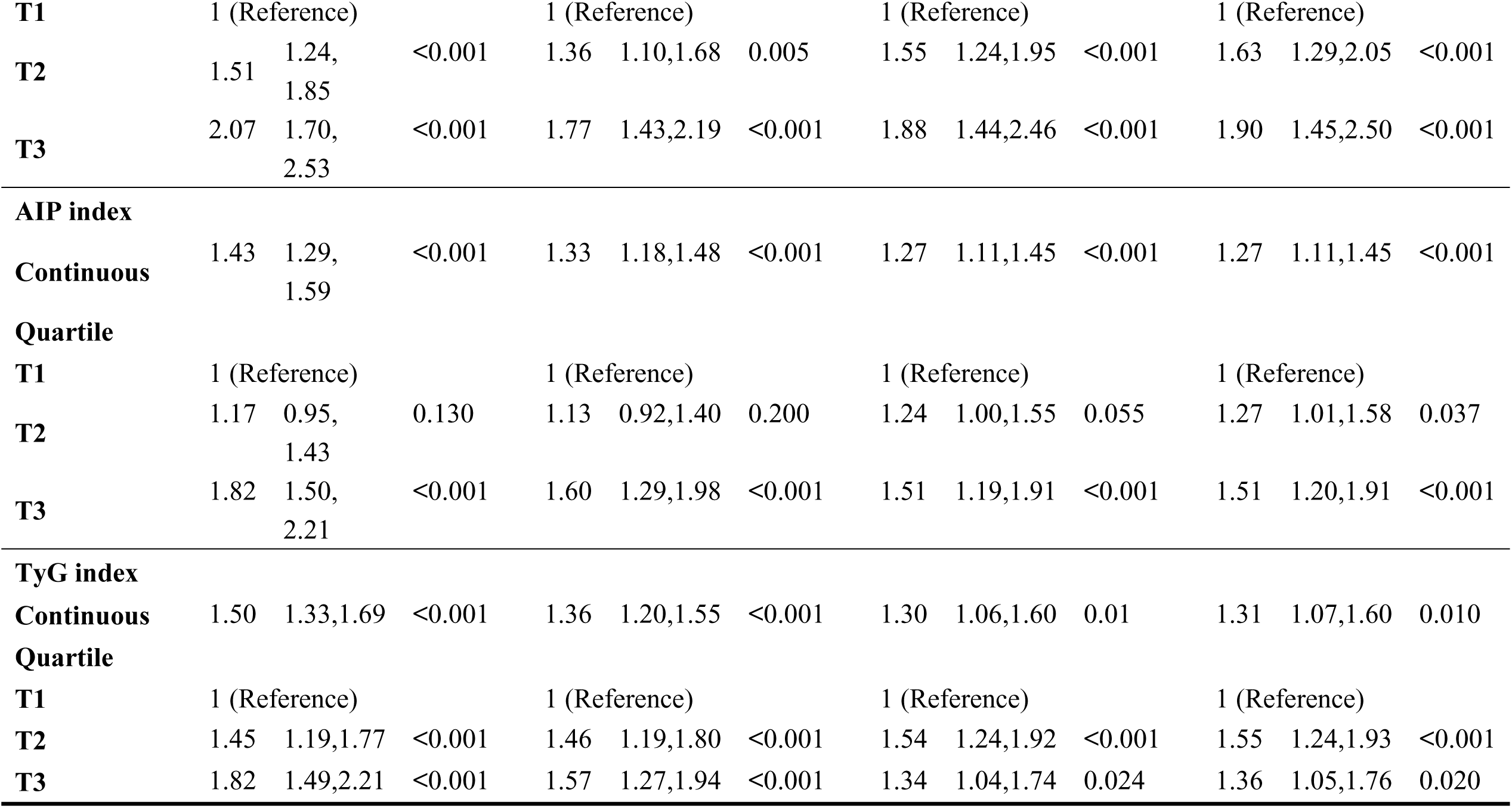
Association of CHG, AIP, and TyG Indices with the Risk of New-Onset Hypertension: Results from Multivariable Logistic Regression Analysis.

### 3.4 Evaluation of the Association Between CHG Index, AIP Index, and TyG Index and New-Onset Hypertension Using RCS Models

In the unadjusted RCS models, a significant positive linear association was observed between the CHG index (Figure 2A) and incident hypertension (overall P < 0.001, nonlinear P = 0.403). Similar linear trends were found for both the AIP and TyG indices (Figures 2B and 2C), with overall P-values < 0.001 and nonlinear P-values of 0.152 and 0.816, respectively. After adjusting for multiple covariates—including sex, age, residence area, marital status, education level, smoking and alcohol consumption habits, baseline SBP and DBP, WBC, HGB, PLT, TC, LDL-C, uric acid, creatinine, cystatin C, and BUN—the association between the CHG index and new-onset hypertension revealed a significant positive nonlinear trend (Figure 2D; overall P < 0.001, nonlinear P = 0.042). In contrast, the associations for the AIP and TyG indices remained linear after full adjustment (Figure 2E and 2F), with overall P-values of 0.002 and 0.022, and nonlinear P-values of 0.849 and 0.292, respectively. The monotonic increasing relationships observed suggest that individuals with higher levels of CHG, AIP, or TyG indices may be at greater risk of developing hypertension during the prehypertensive stage. These findings highlight the potential utility of these metabolic indices in the early identification and risk stratification of individuals at risk for hypertension.

**Fig. 2.**
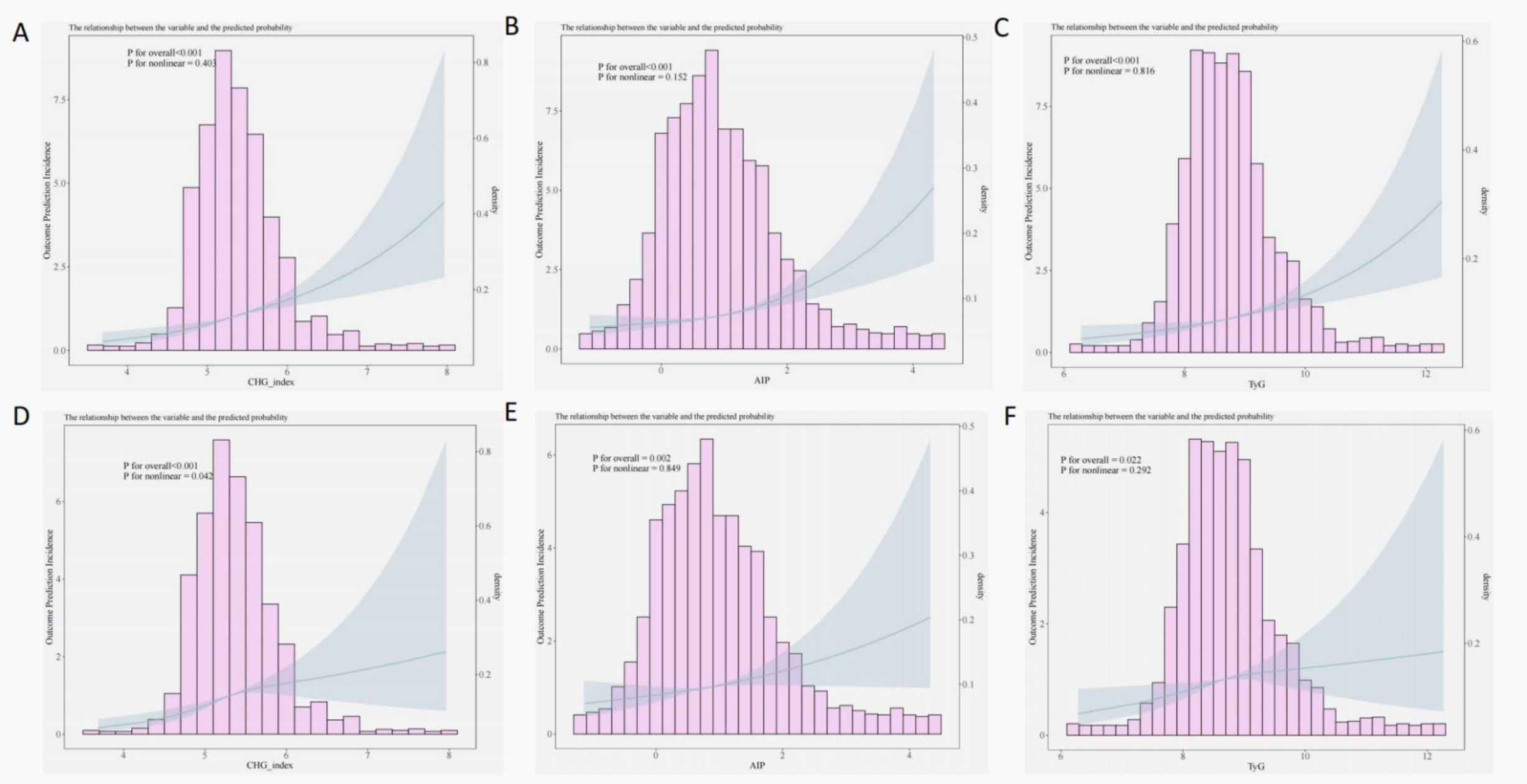
Restricted cubic spline models.

### 3.5 Comparison of CHG, AIP, and TyG Indices in Predicting New-Onset Hypertension After 4-Year Follow-Up in Prehypertensive Individuals

In the unadjusted ROC analysis, the CHG, AIP, and TyG indices showed comparable performance in predicting incident hypertension over a 4-year follow-up period among individuals with prehypertension. Specifically, the AUC was 0.5877 (95% CI: 0.5653–0.6100) for the CHG index, 0.5763 (95% CI: 0.5536–0.5991) for the AIP index, and 0.5727 (95% CI: 0.5501–0.5953) for the TyG index (Figure 3A). These results indicate that, without adjustment, all three indices demonstrated similar predictive abilities for new-onset hypertension, with the CHG index showing a slightly higher AUC than the other two, although the differences were small. However, after multivariable adjustment for potential confounders—including sex, age, residence area, smoking and alcohol consumption habits, marital status, education level, baseline SBP and DBP, WBC, HGB, lipid profiles (TC and LDL-C), uric acid, and other biochemical parameters—the predictive performance of all indices improved significantly. In the fully adjusted model, the AUC for the CHG index reached 0.7010 (95% CI: 0.6801–0.7219), followed by the AIP index at 0.6997 (95% CI: 0.6787–0.7207) and the TyG index at 0.6980 (95% CI: 0.6770–0.7190) (Figure 3B). Notably, the CHG index consistently exhibited the highest predictive accuracy across both the unadjusted and fully adjusted models, suggesting its robustness and stability as a potential predictor of hypertension development in individuals with prehypertension.

**Fig. 3.**
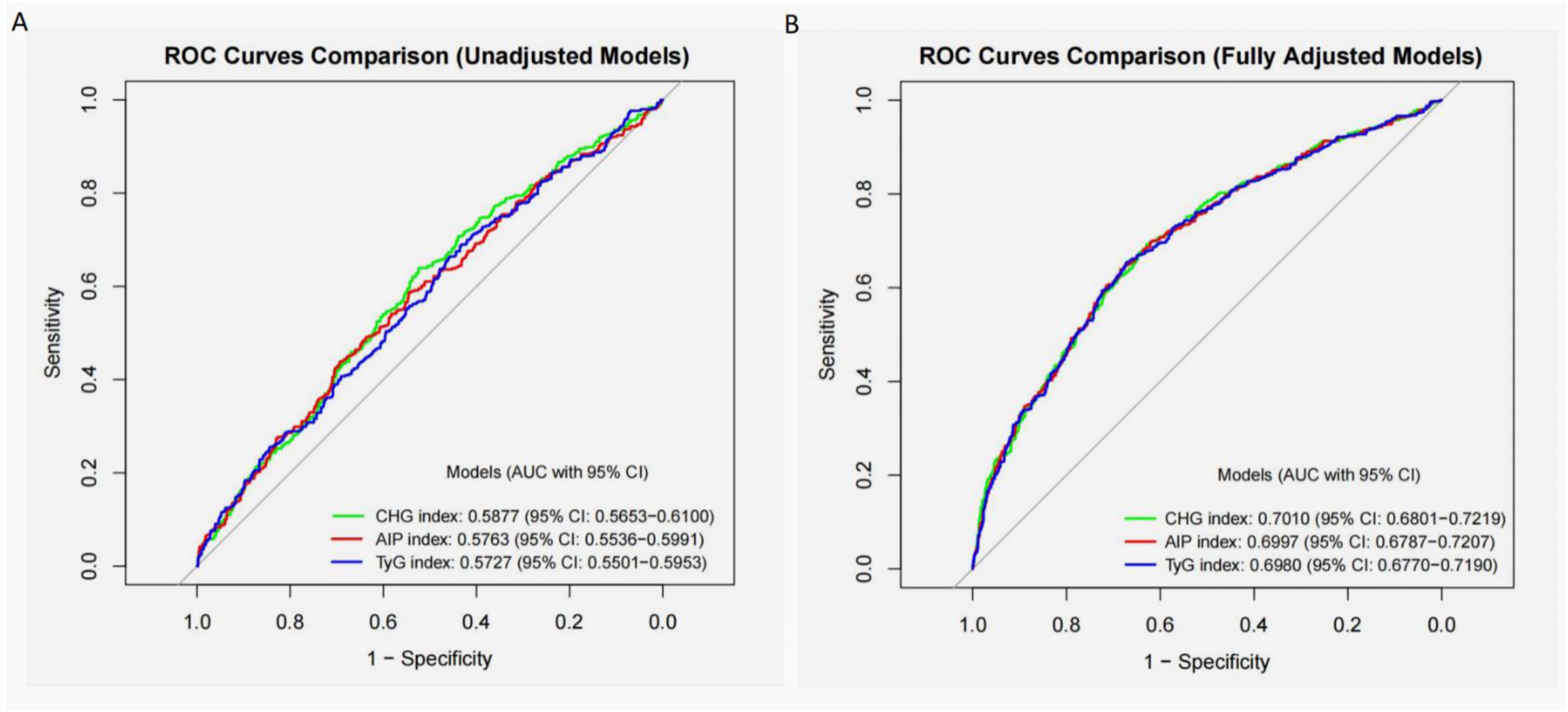
ROC Curve Analysis Comparing the Predictive Performance of CHG, AIP, and TyG Indices for Incident Hypertension in Prehypertensive Individuals: Unadjusted vs. Fully Adjusted Models

### 3.6 Subgroup Analysis of the Relationship Between CHG Index, AIP Index, and TyG Index and New-Onset Hypertension

Subgroup analyses were conducted based on education level, age, marital status, residence area, alcohol consumption, smoking status, BMI, CVD, DM, depression status, and life satisfaction, with adjustment for relevant covariates. Figure 4 presents the results of the subgroup analysis for the association between the CHG index and incident hypertension, revealing stronger associations within specific subpopulations. Specifically, the CHG index showed significantly greater effects among individuals with lower educational attainment (illiterate), those aged 60–70 or ≥70 years, widowed individuals, and those without DM (P < 0.05 for interaction). In contrast, no significant heterogeneity was observed across subgroups defined by sex, residence area, alcohol consumption, BMI, CVD, or depression status (all P-values for interaction > 0.05), indicating a relatively stable association across these characteristics, although the effect was more pronounced in certain high-risk populations. Detailed results of the subgroup analyses for the AIP and TyG indices are presented in Supplementary Figures S1 and S2. The association between the AIP index and new-onset hypertension was not significantly modified by education level, marital status, residence area, alcohol consumption, depression status, or DM (all P-values for interaction > 0.05). However, the association was notably stronger among females, individuals aged ≥70 years, those with a BMI of 18.5–24 kg/m², participants with CVD, and those reporting a life satisfaction score of 2 (P < 0.05 for all; P-values for interaction < 0.05). Similarly, the association between the TyG index and incident hypertension remained consistent across subgroups defined by sex, education level, residence area, alcohol consumption, BMI, DM, or depression status (all P-values for interaction > 0.05). However, a stronger relationship was observed in individuals aged ≥70 years, widowed individuals, those with CVD, and those reporting a life satisfaction score of 2 (P < 0.05 for all; P-values for interaction < 0.05). Overall, while the associations of the CHG, AIP, and TyG indices with new-onset hypertension were more pronounced in certain high-risk subgroups, they remained largely consistent across different population strata. These findings highlight the potential utility of these metabolic indices in identifying and managing individuals at increased risk of developing hypertension, particularly in vulnerable subpopulations.

**Fig. 4.**
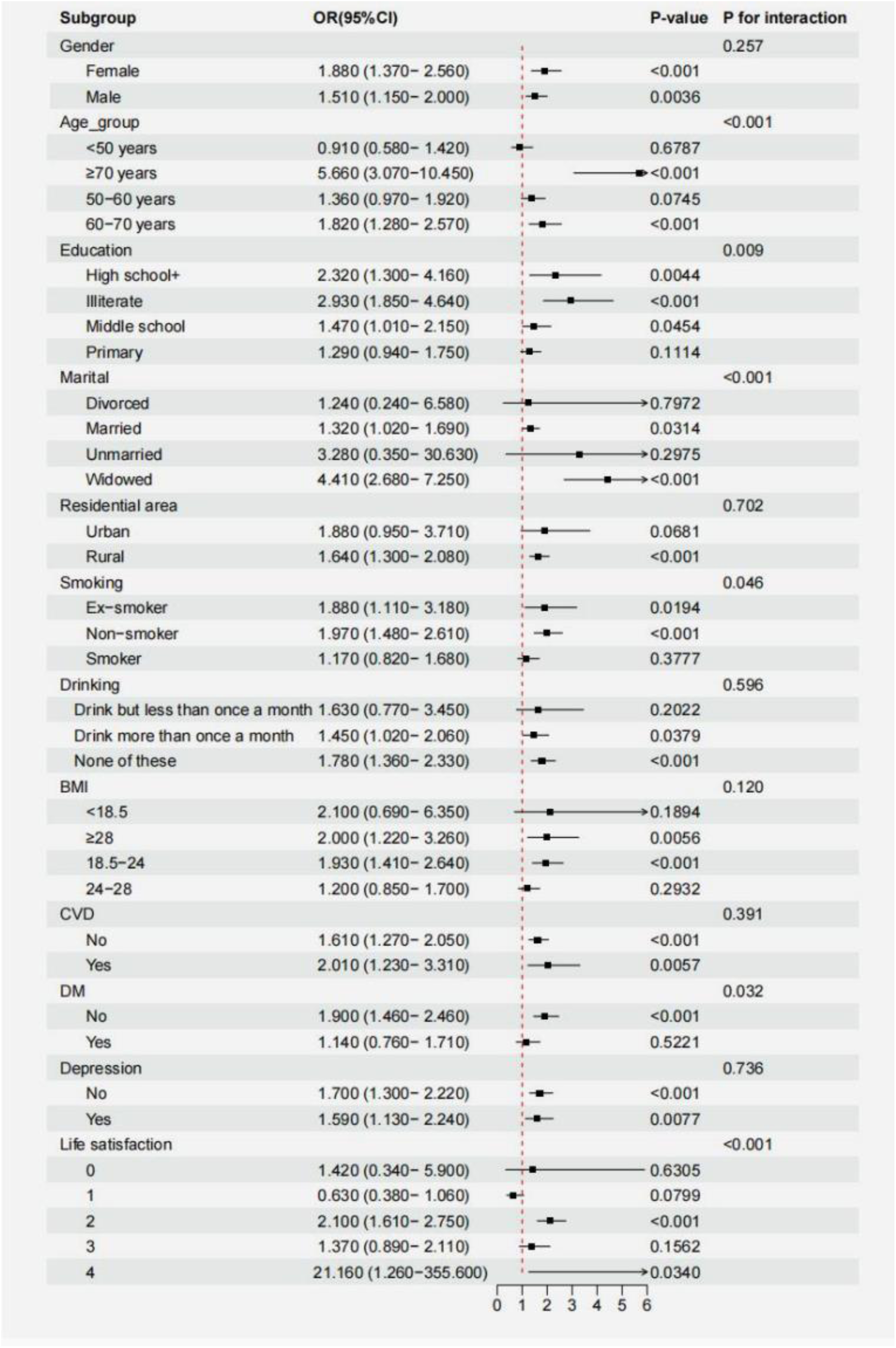
Forest Plot of Multivariable-Adjusted Associations Between CHG Index and Incident Hypertension by Subgroup

## 4. Discussion

To our knowledge, this study represents the first comprehensive evaluation comparing the predictive value of three metabolic indices (CHG, AIP, and TyG) for incident hypertension in prehypertensive individuals. Our analysis of the CHARLS database yielded three key findings: First, all three indices independently predicted hypertension risk after multivariable adjustment, with CHG index demonstrating superior predictive performance (OR: 1.96, 95%CI 1.45-2.66, P<0.001) compared to TyG index (OR: 1.31, 95%CI 1.07-1.60, P=0.010).

Second, restricted cubic spline analyses revealed a significant nonlinear dose-response relationship between CHG index and hypertension risk (P for nonlinear=0.042), contrasting with the linear associations observed for AIP and TyG indices. Third, receiver operating characteristic analysis confirmed CHG index’s discriminative advantage (fully adjusted model AUC=0.7010) over AIP (AUC=0.6997) and TyG (AUC=0.6980) indices. These findings position CHG index as a potentially superior tool for risk stratification in prehypertension management.

This study systematically compared the predictive value of three metabolic indices—CHG, AIP, and TyG—in individuals with prehypertension, providing important evidence for research in this field. First, regarding the AIP, this study provides new evidence to address existing controversies and further expands upon previous findings. Although several prior studies have explored the association between AIP and the risk of hypertension, results have been heterogeneous^14–17,29,30^. In a cross-sectional study involving 15,453 participants with normal glucose levels in Japan, Tan et al. found a positive association between AIP and hypertension risk, particularly among women aged 40–60 years^14^. This finding was consistent with that of Yuan et al., who conducted a cohort study of 3,150 individuals in China and identified 1,054 incident cases of hypertension over a 6-year follow-up period^15^. Prospective studies from Taiwan further indicated that AIP had significant predictive value among individuals aged 40–64 years, but this association weakened in those aged over 65 years, with stronger associations observed in males (OR = 1.51)^29^. These results were similar to findings from a study conducted in Turkey^16^. However, a cross-sectional study in mainland China did not find a significant association between AIP and hypertension in a non-diabetic population, suggesting that population-specific differences may influence the predictive performance of AIP^30^. By employing a more rigorous study design and a larger sample size (n = 2,859), our study provides new evidence to help resolve this controversy. We found that individuals in the highest tertile of AIP (T3) had a significantly increased risk of developing hypertension (OR: 1.51, 95% CI: 1.20–1.91, P < 0.001), supporting the conclusions of most previous studies. Notably, consistent with the findings of Tan et al. and Yuan et al., our subgroup analysis revealed that AIP demonstrated enhanced predictive value in specific subpopulations, including females, individuals aged ≥70 years, those with BMI 18.5–24 and existing CVD, and individuals with a life satisfaction score of 2 (P < 0.05)^14,15^. However, unlike the previously reported male predominance in studies from Taiwan and Turkey, our data showed that AIP had greater predictive power in females (interaction P < 0.05), a discrepancy that may be attributable to differences in the metabolic profiles of the studied populations^16,29^. Moreover, our findings indicate that the predictive value of AIP was not significantly influenced by factors such as education level, marital status, residential area, alcohol consumption, depression, or DM (interaction P > 0.05).

In exploring the association between the TyG index and the risk of hypertension, this study provides novel insights by synthesizing previously conflicting evidence. Similar to AIP, the predictive value of the TyG index has shown significant heterogeneity across studies. Numerous epidemiological studies have confirmed that individuals with prehypertension are at 2–3 times higher risk of progressing to hypertension compared to those with normal blood pressure, a finding highly consistent with the 30.3% progression rate reported in the Framingham Heart Study^31–34^. Against this backdrop, the predictive value of the TyG index—as a surrogate marker for insulin resistance—has remained controversial. A large cross-sectional study by Song et al. supported a positive association between the TyG index and hypertension, whereas Liu et al. found no significant relationship among Chinese individuals with normal body weight^20,21^. This study, based on a large cohort (n = 2,859) and rigorous multivariable adjustment, confirms that individuals in the highest tertile of the TyG index (T3) had a significantly increased risk of incident hypertension (OR: 1.36, 95% CI: 1.05–1.76, P = 0.020), supporting the main findings of Song et al.^21^. Using RCS analysis, we observed a stable positive linear relationship between the TyG index and hypertension risk (overall P = 0.022; nonlinear P = 0.292). The predictive performance of the TyG index, as measured by the area under the receiver operating characteristic curve (AUC = 0.6980, 95% CI: 0.6770–0.7190), was consistent across different models. Notably, in contrast to the null results reported by Liu et al., our subgroup analyses identified important effect modifiers: the TyG index demonstrated enhanced predictive value in high-risk subgroups such as individuals aged ≥70 years (OR = 1.63), widowed individuals, and those with CVD (interaction P < 0.05)^20^. This may be attributed to the more severe metabolic disturbances commonly observed in these populations. Importantly, the TyG index maintained consistent predictive performance across conventional stratification variables such as sex and BMI (interaction P > 0.05), making it a reliable tool in clinical practice.

Most importantly, this study represents the first systematic validation of the novel metabolic index—CHG—in predicting hypertension, offering a breakthrough tool for the early identification of metabolic syndrome-related hypertension. Originally developed by Mansoori et al. for DM diagnosis, the CHG index demonstrated several advantages over traditional metabolic markers such as AIP and TyG in this study^24^. In the fully adjusted model, the CHG index exhibited superior predictive performance (AUC = 0.7010, 95% CI: 0.6801–0.7219), outperforming both AIP (AUC = 0.6997) and the TyG index (AUC = 0.6980), and maintaining the best stability across models. Particularly noteworthy was the unique nonlinear dose–response relationship revealed by RCS analysis (P nonlinearity = 0.042), an intriguing parallel to the findings of Mansoori et al. in DM research, suggesting that the CHG index may more accurately reflect the complex pathophysiological processes linking metabolic disturbance and elevated blood pressure^24^. Unlike AIP and TyG, the CHG index demonstrated strong generalizability across populations, with its predictive value unaffected by common confounding factors such as sex, residential area, or alcohol consumption (all interaction P > 0.05). However, in specific high-risk subgroups—including individuals over 60 years of age, widowed individuals, and those without DM—the CHG index showed even stronger associations (P < 0.05), highlighting its potential utility in targeted prevention strategies.

The associations observed in this study between three metabolic indices—CHG index, AIP, and TyG index—and the incidence of hypertension among individuals with prehypertension may be jointly mediated through a series of complex pathophysiological mechanisms. First, all these indices reflect core features of metabolic syndrome, with insulin resistance being one of the key underlying factors^35,36^. Insulin resistance contributes directly to elevated blood pressure by activating the renin-angiotensin system and inducing endothelial dysfunction^36,37^. In addition, lipid abnormalities, particularly elevated TG and reduced HDL-C, impair the bioavailability of nitric oxide, thereby affecting vascular relaxation function^38^. Meanwhile, visceral fat accumulation not only increases the release of inflammatory cytokines^35,39^, but also leads to dysregulation of the autonomic nervous system, further exacerbating the formation of atherosclerotic plaques and reducing arterial wall compliance, ultimately promoting the elevation of blood pressure^36,37,40,41^. Beyond these shared mechanisms, each metabolic index also has distinct mechanistic pathways. For instance, AIP primarily exerts its negative effects on vascular function by promoting the formation of small, dense LDL-C particles and increasing oxidative stress levels^42–45^. The TyG index focuses specifically on adipokine imbalance associated with central obesity, a state that plays an important role in the development of hypertension^46,47^. In contrast, the CHG index integrates the ratio of total cholesterol to HDL-C and FBG, enabling it to comprehensively reflect both impaired cholesterol reverse transport and abnormal glucose metabolism, thus offering broader predictive value^48,49^. Notably, the reduction in HDL-C levels not only decreases the production of endothelium-derived nitric oxide, leading to vascular dysfunction, but also promotes increased arterial stiffness via oxidative stress pathways, both of which are major driving forces in the development of hypertension^40,41,50^. In summary, these findings provide a pathophysiological basis for understanding the differences in predictive performance among various metabolic indices, and help explain why the CHG index—which integrates multiple mechanistic pathways—demonstrates the best predictive performance.

This study demonstrates several notable strengths. It is the first systematic evaluation of the CHG index for predicting the risk of incident hypertension among individuals with prehypertension, and directly compares its predictive performance with two well-established metabolic indices—AIP and TyG. This comparative analysis provides novel insights into risk assessment for new-onset hypertension. The study employed a comprehensive set of statistical methods, including logistic regression, RCS analysis, and ROC curve modeling, thereby establishing a robust and multi-dimensional evaluation framework. These diverse analytical approaches not only enhance the methodological rigor of the study but also strengthen the validity and interpretability of the results. Furthermore, by utilizing data from a large cohort database, the study effectively minimizes selection bias and ensures sample representativeness. We validated the CHG index as a reliable risk prediction tool and particularly demonstrated its enhanced predictive value in high-risk subgroups, such as individuals aged ≥70 years, widowed individuals, and those with CVD. Given the global prevalence and high incidence of prehypertension, this research carries significant clinical implications and public health relevance. The CHG index is simple to calculate and not constrained by time or cost, making it highly suitable for large-scale population screening. As a risk stratification tool for individuals with prehypertension, it enables early identification and timely intervention for incident hypertension, facilitating preventive strategies that can reduce disease progression and complications, improve quality of life, and alleviate socioeconomic burdens. Therefore, this study not only provides clinicians with a valuable new reference metric but also offers scientific evidence to support the development of public health policies aimed at targeted interventions in broader populations.

This study has several limitations that should be acknowledged. First, due to the observational nature of the study design, we cannot establish causal relationships between the CHG index, AIP, TyG index, and the incidence of hypertension. Although we adjusted for multiple potential confounders in our analyses, residual or unmeasured confounding factors—such as dietary habits, physical activity levels, lifestyle behaviors, and genetic background—may still influence the observed associations. In particular, diet plays a significant role in modulating both metabolic profiles and blood pressure levels^45^. High carbohydrate and high-fat diets can markedly affect these indices and blood pressure; however, this study did not account for dietary patterns or assess their potential impact on our findings. Second, key variables including the CHG index, AIP, and TyG index were only measured at baseline, lacking dynamic assessments of changes over time. Therefore, we were unable to evaluate whether fluctuations in these metabolic markers are associated with changes in blood pressure levels.Third, the data used in this study were derived from the CHARLS database, which is nationally representative but primarily focuses on individuals aged 45 years and older. This limits the generalizability of our findings to younger populations. Additionally, some data were collected via self-reported questionnaires, which may introduce information bias. For example, self-reported disease histories may lead to underreporting or misclassification of certain conditions, potentially affecting the accuracy of our results. Moreover, although we adjusted for various demographic and biochemical covariates, the potential influence of underlying comorbidities or other unknown factors on the outcomes cannot be completely ruled out. Future population-based prospective studies are warranted to further elucidate the specific mechanisms linking these metabolic indices with the development of hypertension and to identify potential intervention targets. To improve upon the current study design, future research should consider conducting prospective cohort studies or interventional trials with regular monitoring of changes in metabolic indices such as CHG, AIP, and TyG, and their associations with blood pressure progression. Furthermore, interventional studies targeting lifestyle modifications—such as dietary optimization and structured exercise programs—or pharmacological approaches aimed at reducing CHG levels could be implemented to determine whether lowering CHG is associated with a reduced risk of incident hypertension. Such investigations would help elucidate the potential causal role of CHG in hypertension development and further validate its preventive utility. In addition, future research should explore other candidate biomarkers and their potential interactions with the CHG index, as well as evaluate the performance of the CHG index across different stages of prehypertension as individuals transition to established hypertension.

## 5. Conclusion

This study shows that the CHG index, AIP index, and TyG index are all significantly associated with an increased risk of developing new hypertension in individuals with prehypertension. Among these, the CHG index performs best, demonstrating a significant positive nonlinear trend and the highest predictive power. This is particularly evident in high-risk subgroups such as those aged 70 or older, widowed individuals, or those with CVD. These findings indicate that the CHG index, as a new and important tool, has significant advantages in early identification and management of the risk of prehypertensive individuals progressing to hypertension. This simple and easily accessible indicator can help advance the development of early prevention and personalized management strategies for hypertension.

## Data Availability

This study utilized data from the China Health and Retirement Longitudinal Study, which is accessible to researchers at http://charls.pku.edu.cn/.

## Acknowledgment

We are grateful for the access to the China Health and Retirement Longitudinal Study database, which significantly enriched our research.

## Author Contributions

Mengmeng Wang, having full access to the dataset, was responsible for ensuring the accuracy and completeness of the analyses conducted. As co-first authors, Mengmeng Wang, Zhankui Du, and Tianqi Teng made equivalent contributions to the execution of the study, including data collection, analysis, interpretation, and the preparation of the initial draft. Jiachao Xu, Zihan Dong, and Qingying Jiao participated in the data collection and analysis processes. The conceptualization of the study and the review of the manuscript were undertaken by Haichu Yu, Ning Zhang, and Mengmeng Wang. All authors have read and approved the final manuscript.

## Ethics Statement

Not applicable.

## References

1. Egan BM, Stevens-Fabry S. Prehypertension--prevalence, health risks, and management strategies. Nat Rev Cardiol. May 2015;12(5):289–300. doi:10.1038/nrcardio.2015.17

2. Heidenreich PA, Trogdon JG, Khavjou OA, et al. Forecasting the future of cardiovascular disease in the United States: a policy statement from the American Heart Association. Circulation. Mar 1 2011;123(8):933–44. doi:10.1161/CIR.0b013e31820a55f5

3. Gu D, Chen J, Wu X, et al. Prehypertension and risk of cardiovascular disease in Chinese adults. J Hypertens. Apr 2009;27(4):721–9. doi:10.1097/HJH.0b013e328323ad89

4. Choi KM, Park HS, Han JH, et al. Prevalence of prehypertension and hypertension in a Korean population: Korean National Health and Nutrition Survey 2001. J Hypertens. Aug 2006;24(8):1515–21. doi:10.1097/01.hjh.0000239286.02389.0f

5. Huang Y, Wang S, Cai X, et al. Prehypertension and incidence of cardiovascular disease: a meta-analysis. BMC Med. Aug 2 2013;11:177. doi:10.1186/1741-7015-11-177

6. Chobanian AV, Bakris GL, Black HR, et al. Seventh report of the Joint National Committee on Prevention, Detection, Evaluation, and Treatment of High Blood Pressure. Hypertension. Dec 2003;42(6):1206–52. doi:10.1161/01.HYP.0000107251.49515.c2

7. Global burden and strength of evidence for 88 risk factors in 204 countries and 811 subnational locations, 1990-2021: a systematic analysis for the Global Burden of Disease Study 2021. Lancet. May 18 2024;403(10440):2162–2203. doi:10.1016/s0140-6736(24)00933-4

8. Lawes CM, Bennett DA, Feigin VL, Rodgers A. Blood pressure and stroke: an overview of published reviews. Stroke. Mar 2004;35(3):776–85. doi:10.1161/01.Str.0000116869.64771.5a

9. Lewington S, Clarke R, Qizilbash N, Peto R, Collins R. Age-specific relevance of usual blood pressure to vascular mortality: a meta-analysis of individual data for one million adults in 61 prospective studies. Lancet. Dec 14 2002;360(9349):1903–13. doi:10.1016/s0140-6736(02)11911-8

10. Dobiásová M, Frohlich J. [The new atherogenic plasma index reflects the triglyceride and HDL-cholesterol ratio, the lipoprotein particle size and the cholesterol esterification rate: changes during lipanor therapy]. Vnitr Lek. Mar 2000;46(3):152–6. Nový aterogenní index plazmy (AIP) odpovídá pomĕru triglyceridů a HDL-cholesterolu, velikosti cástic lipoproteinů a esterifikacní rychlosti cholesterolu: zmĕny po lécbĕ lipanorem.

11. Won KB, Jang MH, Park EJ, et al. Atherogenic index of plasma and the risk of advanced subclinical coronary artery disease beyond traditional risk factors: An observational cohort study. Clin Cardiol. Dec 2020;43(12):1398–1404. doi:10.1002/clc.23450

12. Liu H, Liu K, Pei L, et al. Atherogenic Index of Plasma Predicts Outcomes in Acute Ischemic Stroke. Front Neurol. 2021;12:741754. doi:10.3389/fneur.2021.741754

13. Kim SH, Cho YK, Kim YJ, et al. Association of the atherogenic index of plasma with cardiovascular risk beyond the traditional risk factors: a nationwide population-based cohort study. Cardiovasc Diabetol. May 22 2022;21(1):81. doi:10.1186/s12933-022-01522-8

14. Tan M, Zhang Y, Jin L, et al. Association between atherogenic index of plasma and prehypertension or hypertension among normoglycemia subjects in a Japan population: a cross-sectional study. Lipids Health Dis. Jun 29 2023;22(1):87. doi:10.1186/s12944-023-01853-9

15. Yuan Y, Shi J, Sun W, Kong X. The positive association between the atherogenic index of plasma and the risk of new-onset hypertension: a nationwide cohort study in China. Clin Exp Hypertens. Dec 31 2024;46(1):2303999. doi:10.1080/10641963.2024.2303999

16. Onat A, Can G, Kaya H, Hergenç G. “Atherogenic index of plasma” (log10 triglyceride/high-density lipoprotein-cholesterol) predicts high blood pressure, diabetes, and vascular events. J Clin Lipidol. Mar-Apr 2010;4(2):89–98. doi:10.1016/j.jacl.2010.02.005

17. Choudhary MK, Eräranta A, Koskela J, et al. Atherogenic index of plasma is related to arterial stiffness but not to blood pressure in normotensive and never-treated hypertensive subjects. Blood Press. Jun 2019;28(3):157–167. doi:10.1080/08037051.2019.1583060

18. Sun Y, Ji H, Sun W, An X, Lian F. Triglyceride glucose (TyG) index: A promising biomarker for diagnosis and treatment of different diseases. Eur J Intern Med. Jan 2025;131:3–14. doi:10.1016/j.ejim.2024.08.026

19. Zhang F, Zhang Y, Guo Z, et al. The association of triglyceride and glucose index, and triglyceride to high-density lipoprotein cholesterol ratio with prehypertension and hypertension in normoglycemic subjects: A large cross-sectional population study. J Clin Hypertens (Greenwich). Jul 2021;23(7):1405–1412. doi:10.1111/jch.14305

20. Liu XZ, Fan J, Pan SJ. METS-IR, a novel simple insulin resistance indexes, is associated with hypertension in normal-weight Chinese adults. J Clin Hypertens (Greenwich). Aug 2019;21(8):1075–1081. doi:10.1111/jch.13591

21. Jian S, Su-Mei N, Xue C, Jie Z, Xue-Sen W. Association and interaction between triglyceride-glucose index and obesity on risk of hypertension in middle-aged and elderly adults. Clin Exp Hypertens. 2017;39(8):732–739. doi:10.1080/10641963.2017.1324477

22. Morales-Gurrola G, Simental-Mendía LE, Castellanos-Juárez FX, Salas-Pacheco JM, Guerrero-Romero F. The triglycerides and glucose index is associated with cardiovascular risk factors in metabolically obese normal-weight subjects. J Endocrinol Invest. Jul 2020;43(7):995–1000. doi:10.1007/s40618-020-01184-x

23. Wang K, He G, Zhang Y, et al. Association of triglyceride-glucose index and its interaction with obesity on hypertension risk in Chinese: a population-based study. J Hum Hypertens. Mar 2021;35(3):232–239. doi:10.1038/s41371-020-0326-4

24. Mansoori A, Nosrati M, Dorchin M, et al. A novel index for diagnosis of type 2 diabetes mellitus: Cholesterol, High density lipoprotein, and Glucose (CHG) index. J Diabetes Investig. Feb 2025;16(2):309–314. doi:10.1111/jdi.14343

25. Zhao Y, Hu Y, Smith JP, Strauss J, Yang G. Cohort profile: the China Health and Retirement Longitudinal Study (CHARLS). Int J Epidemiol. Feb 2014;43(1):61–8. doi:10.1093/ije/dys203

26. Gong J, Wang G, Wang Y, et al. Nowcasting and forecasting the care needs of the older population in China: analysis of data from the China Health and Retirement Longitudinal Study (CHARLS). Lancet Public Health. Dec 2022;7(12):e1005–e1013. doi:10.1016/s2468-2667(22)00203-1

27. Chen X, Crimmins E, Hu PP, et al. Venous Blood-Based Biomarkers in the China Health and Retirement Longitudinal Study: Rationale, Design, and Results From the 2015 Wave. Am J Epidemiol. Nov 1 2019;188(11):1871–1877. doi:10.1093/aje/kwz170

28. Luo H, Li J, Zhang Q, et al. Obesity and the onset of depressive symptoms among middle-aged and older adults in China: evidence from the CHARLS. BMC Public Health. Jul 24 2018;18(1):909. doi:10.1186/s12889-018-5834-6

29. Li YW, Kao TW, Chang PK, Chen WL, Wu LW. Atherogenic index of plasma as predictors for metabolic syndrome, hypertension and diabetes mellitus in Taiwan citizens: a 9-year longitudinal study. Sci Rep. May 10 2021;11(1):9900. doi:10.1038/s41598-021-89307-z

30. Cheng W, Zhuang J, Chen S. Dyslipidemia and the Prevalence of Hypertension: A Cross-Sectional Study Based on Chinese Adults Without Type 2 Diabetes Mellitus. Front Cardiovasc Med. 2022;9:938363. doi:10.3389/fcvm.2022.938363

31. Lüders S, Schrader J, Berger J, et al. The PHARAO study: prevention of hypertension with the angiotensin-converting enzyme inhibitor ramipril in patients with high-normal blood pressure: a prospective, randomized, controlled prevention trial of the German Hypertension League. J Hypertens. Jul 2008;26(7):1487–96. doi:10.1097/HJH.0b013e3282ff8864

32. Faselis C, Doumas M, Kokkinos JP, et al. Exercise capacity and progression from prehypertension to hypertension. Hypertension. Aug 2012;60(2):333–8. doi:10.1161/hypertensionaha.112.196493

33. Selassie A, Wagner CS, Laken ML, Ferguson ML, Ferdinand KC, Egan BM. Progression is accelerated from prehypertension to hypertension in blacks. Hypertension. Oct 2011;58(4):579–87. doi:10.1161/hypertensionaha.111.177410

34. Vasan RS, Larson MG, Leip EP, Kannel WB, Levy D. Assessment of frequency of progression to hypertension in non-hypertensive participants in the Framingham Heart Study: a cohort study. Lancet. Nov 17 2001;358(9294):1682–6. doi:10.1016/s0140-6736(01)06710-1

35. Sironi AM, Gastaldelli A, Mari A, et al. Visceral fat in hypertension: influence on insulin resistance and beta-cell function. Hypertension. Aug 2004;44(2):127–33. doi:10.1161/01.HYP.0000137982.10191.0a

36. DeMarco VG, Aroor AR, Sowers JR. The pathophysiology of hypertension in patients with obesity. Nat Rev Endocrinol. Jun 2014;10(6):364–76. doi:10.1038/nrendo.2014.44

37. Oparil S, Zaman MA, Calhoun DA. Pathogenesis of hypertension. Ann Intern Med. Nov 4 2003;139(9):761–76. doi:10.7326/0003-4819-139-9-200311040-00011

38. McGill JB, Haffner S, Rees TJ, Sowers JR, Tershakovec AM, Weber M. Progress and controversies: treating obesity and insulin resistance in the context of hypertension. J Clin Hypertens (Greenwich). Jan 2009;11(1):36–41. doi:10.1111/j.1751-7176.2008.00065.x

39. Morales-Villegas E. Dyslipidemia, Hypertension and Diabetes Metaflammation. A Unique Mechanism for 3 Risk Factors. Curr Hypertens Rev. Jul 1 2014;

40. Jonas K, Magoń W, Podolec P, Kopeć G. Triglyceride-to-High-Density Lipoprotein Cholesterol Ratio and Systemic Inflammation in Patients with Idiopathic Pulmonary Arterial Hypertension. Med Sci Monit. Jan 26 2019;25:746–753. doi:10.12659/msm.912766

41. Angeli F, Reboldi G, Gentile G, Verdecchia P. The emerging role of high-density lipoprotein cholesterol in hypertension trials. J Hypertens. Mar 2009;27(3):458–60. doi:10.1097/HJH.0b013e3283232a59

42. Austin MA, Breslow JL, Hennekens CH, Buring JE, Willett WC, Krauss RM. Low-density lipoprotein subclass patterns and risk of myocardial infarction. Jama. Oct 7 1988;260(13):1917–21.

43. Anber V, Griffin BA, McConnell M, Packard CJ, Shepherd J. Influence of plasma lipid and LDL-subfraction profile on the interaction between low density lipoprotein with human arterial wall proteoglycans. Atherosclerosis. Aug 2 1996;124(2):261–71. doi:10.1016/0021-9150(96)05842-x

44. Zambon A, Austin MA, Brown BG, Hokanson JE, Brunzell JD. Effect of hepatic lipase on LDL in normal men and those with coronary artery disease. Arterioscler Thromb. Feb 1993;13(2):147–53. doi:10.1161/01.atv.13.2.147

45. Zhao T, Wu K, Hogstrand C, et al. Lipophagy mediated carbohydrate-induced changes of lipid metabolism via oxidative stress, endoplasmic reticulum (ER) stress and ChREBP/PPARγ pathways. Cell Mol Life Sci. May 2020;77(10):1987–2003. doi:10.1007/s00018-019-03263-6

46. Bühler FR, Tkachuk VA, Hahn AW, Resink TJ. Low- and high-density lipoproteins as hormonal regulators of platelet, vascular endothelial and smooth muscle cell interactions: relevance to hypertension. J Hypertens Suppl. Dec 1991;9(6):S28–36.

47. Galle J, Ochslen M, Schollmeyer P, Wanner C. Oxidized lipoproteins inhibit endothelium-dependent vasodilation. Effects of pressure and high-density lipoprotein. Hypertension. May 1994;23(5):556–64. doi:10.1161/01.hyp.23.5.556

48. Mineo C, Deguchi H, Griffin JH, Shaul PW. Endothelial and antithrombotic actions of HDL. Circ Res. Jun 9 2006;98(11):1352–64. doi:10.1161/01.Res.0000225982.01988.93

49. Barter PJ, Nicholls S, Rye KA, Anantharamaiah GM, Navab M, Fogelman AM. Antiinflammatory properties of HDL. Circ Res. Oct 15 2004;95(8):764–72. doi:10.1161/01.Res.0000146094.59640.13

50. Hayakawa H, Raij L. Relationship between hypercholesterolaemia, endothelial dysfunction and hypertension. J Hypertens. May 1999;17(5):611–9. doi:10.1097/00004872-199917050-00004

